# Consensus Guideline for the Management of Colorectal Cancer with Peritoneal Metastases

**DOI:** 10.1101/2024.05.07.24305476

**Authors:** PSM Writing Group, PSM Consortium Group, Kiran K. Turaga

## Abstract

**Background:** The peritoneum is a common site of metastases from colorectal cancer (CRC), yet controversy exists regarding optimal treatment strategies. These guidelines describe the results of a national consensus addressing the management of CRC with peritoneal metastases (CRC-PM).

**Methods:** An update of the 2018 Chicago Consensus Guidelines was conducted using a modified Delphi technique. Two rounds of voting were performed to assess agreement levels on two clinical management pathways regarding synchronous and metachronous CRC-PM. Supporting evidence was evaluated via rapid literature reviews.

**Results:** The overall level of evidence was low in existing literature. Of 145 participants in the first round, 136 (96.8%) responded in the second round. Over 90% consensus was achieved in most pathway blocks. For both pathways, early referral to a peritoneal surface malignancy (PSM) center should be made for patients with CRC-PM. For the synchronous pathway, upfront cytoreductive surgery was de-emphasized in favor of systemic therapy. For the metachronous pathway, risk stratification via clinical and pathologic features was revised. For both pathways, surveillance strategies were added, including only a weak recommendation for circulating tumor DNA (ctDNA) testing given limited evidence of its utility in detecting and monitoring PM.

**Conclusion:** The consensus-driven clinical pathways provide valuable guidance for the management of CRC-PM. There remains a need for high-quality evidence and prospective multicenter trials in this domain.

**SYNOPSIS:** We developed two consensus-driven clinical pathways for the management of colorectal cancer with peritoneal metastases (CRC-PM), using a modified Delphi approach. Rapid reviews evaluating the optimal systemic therapy and the role of plasma-based liquid-biopsy for CRC-PM were conducted.

## INTRODUCTION

Colorectal cancer (CRC) is the third most prevalent malignancy globally and the second leading cause of cancer-related mortality, with 1.8 million new cases diagnosed annually worldwide [1, 2]. Notably, there is a concerning trend in the rise of early-onset CRC, often presenting with advanced disease in younger patients [3, 4]. Peritoneal metastases (PM) develop in approximately 5-15% of patients with CRC within their disease course. About half of these cases present synchronously with an intact primary tumor and the other half present metachronously in the setting of relapse [1, 5, 6]. However, the incidence of PM might be underestimated as not all high-risk patients undergo diagnostic laparoscopy; this is evidenced by the frequency of PM noted at autopsy [7, 8]. CRC-PM has a worse prognosis compared to other metastatic sites and is associated with malnutrition and bowel obstruction and other complications [9]. As highlighted in a systematic review of clinical trials regarding systemic therapies for metastatic CRC, patients with PM face a poor prognosis with median overall survival of approximately 16 months [10].

Multiple studies, including the PRODIGE 7 trial, have shown promise with cytoreductive surgery (CRS) for CRC-PM, with median overall survival exceeding over 40 months, and some studies have shown benefit with the addition of intraperitoneal chemotherapy (IPCT) compared to systemic chemotherapy alone [11–15]. Yet, several controversies exist regarding management strategies, including the utility of IPCT in therapeutic and prophylactic treatment strategies, optimal sequencing and regimens of systemic therapies, and surveillance modalities. The exclusion of patients with PM from large clinical trials, likely because of the challenges with using the Response Evaluation Criteria in Solid Tumors to assess disease response, precludes a better understanding of these questions [7].

Given the scarcity of evidence guiding treatment decisions, limited standardized pathways exist for managing CRC-PM. This paper builds upon the 2018 Chicago Consensus Guidelines on the management of colorectal metastases and reports a multidisciplinary consensus aimed at outlining the clinical management of synchronous and metachronous CRC-PM [16, 17].

## METHODS

This initiative was part of a national multidisciplinary consortium group process aimed at streamlining guidelines for the care of patients with peritoneal surface malignancies (PSM). The consensus and rapid review methodology has been described in detail in a separate manuscript (submitted) [18]. Major components are summarized below.

### Consensus Group Structure

In brief, the Colorectal Disease Working Group (CDWG) consisted of eight experts (AS, CE, DL, JS, KR, MF, US, JB). A team of 12 trainees, including eight surgical residents (KS, FG, DS, JW, JB, SC, LS, EP), two surgical oncology fellows (MW and NB), and two research fellows (VB, MW) conducted the rapid reviews. Two core group trainee members coordinated the effort (KS and FG).

### Modified Delphi Process

A modified Delphi method with two rounds of voting was employed to gather feedback regarding the clinical management pathways following preliminary synthesis of major updates since 2018. Experts rated their agreement levels on a five-point Likert scale via a Qualtrics questionnaire. A 75% consensus threshold was set and blocks with below 90% agreement underwent further review. Simultaneously, two summary tables outlining first-line systemic and regional therapies for CRC-PM was generated by the CDWG, with directed guidance from the medical oncologist in the working group. These tables were then included in the modified Delphi Round 2 survey for general feedback from the entire PSM consortium.

### Rapid Review of the Literature

The two key questions (KQ) were selected by the CDWG. A MEDLINE search via PubMed between January 2000 and August 2023 was performed for these two key questions.

*Key Question 1*. In patients with CRC-PM undergoing CRS, what are the optimal sequences and regimens of systemic therapy (neoadjuvant, adjuvant, perioperative)?

*Key Question 2*. In patients with CRC-PM, does plasma-based liquid biopsy offer better sensitivity, specificity, and lead time therapy compared with standard surveillance modalities in

a. detecting recurrence following CRS?
b. evaluating response to systemic therapies?

Search strategies were developed and reviewed by a medical librarian specialist (Supplementary Tables 3-5), and the review protocols were pre-registered in PROSPERO (CRD42023471072 and CRD420234778690). The Covidence platform facilitated title and abstract screening, full-text review, and data extraction. Quality assessment was performed using the Newcastle-Ottawa Scale for Key Question 1 and the Quality Assessment of Diagnostic Accuracy Studies Version 2 (QUADAS-2) tool for Key Question 2a and Key Question 2b [19–23]. Articles were screened by two reviewers, and conflicts were resolved by the trainee leads in the CDWG. The review was conducted in alignment with recommendations from the Cochrane Rapid Review Methods Groups and reported in line with the Preferred Reporting Items for Systematic reviews and Meta-Analyses (PRISMA) 2020 guidelines [24, 25].

### External/Patient Perspectives

Members of the Peritoneal Surface Oncology Group International (PSOGI, http://www.psogi.com/) Executive Council were invited to appraise the second version of the two pathways. Their comments were consolidated to evaluate alignment with global practices regarding the management of CRC-PM. Additionally, patients and caregivers from the COLONTOWN support group (https://colontown.org/) reviewed the treatment pathways and offered insights regarding clinical trial enrollment, research outcomes, and available resources for patients with CRC-PM.

## RESULTS

Four pathways were initially proposed: 1) synchronous PM, 2) metachronous PM, 3) prophylactic IPCT for locally advanced CRC, and 4) recurrent CRC-PM post-CRS. However, due to insufficient data, guidelines for prophylactic IPCT and recurrent CRC-PM were not established. Hence, the focus of the current consensus and reviews is on synchronous and metachronous CRC-PM.

### Pathways and Rapid Reviews

In all, 145 experts voted in the first Delphi round, of which 136 (93.8%) responded in the second round. Of survey respondents, 101 (69.7%) were surgical oncologists, 25 (17.2%) were medical oncologists, 12 (8.3%) were pathologists, and 7 (4.8%) belonged to other specialties. Given the low quality of existing evidence in the literature, recommendations were based primarily on expert opinion. The synchronous and metachronous CRC-PM pathways were divided into 11 blocks (Figure 1) and 10 blocks (Figure 2), respectively.

**Figure 1.** Clinical pathway for the management of colorectal cancer with synchronous peritoneal metastases.

**Figure 2.** Clinical pathway for the management of colorectal cancer with metachronous peritoneal metastases.

The rapid reviews cumulatively revealed 2888 abstracts, of which 368 full texts were reviewed. Thirty-four studies were ultimately included for data extraction and quality assessment and cited in relevant sections of the document (PRISMA flow diagrams, Supplementary Figures 1-3).

### Summary of Major Changes

Building upon the 2018 Chicago Consensus Guidelines, the current approach involves a more stringent consensus and review methodology while engaging a larger spectrum of experts and patient advocates [16, 17]. For both pathways, early referral to a PSM center was stressed. For the synchronous pathway, upfront CRS ± IPCT should only be considered in highly select patients, with systemic therapy being the preferred initial treatment. For the metachronous pathway, risk stratification via clinical and pathologic features was revised by considering right-sided tumors and signet ring cell histology as high-risk features and removing younger age as a low-risk feature. For both pathways, repeat CRS ± IPCT can be considered in appropriately selected patients if recurrence is detected after initial CRS ± IPCT. For both pathways, surveillance recommendations were added, which included only a weak recommendation for ctDNA testing given limited evidence of its accuracy for monitoring PM.

### Colorectal Cancer with Synchronous Peritoneal Metastases Pathway (Figure 1, Table 1a, Table 1b)

#### Block 1: Initial Management and Preoperative Considerations

**(% Agreement:** Round 1 = 98%, Round 2 = 99%)

**Table 1.** Modified Delphi Agreement Table.

A cornerstone of the initial management of CRC-PM is early referral to a PSM center. Centralization at high-volume centers is crucial due to the steep institutional learning curve associated with CRS ± IPCT [26]. It is associated with reduced postoperative morbidity and improved oncologic outcomes [27].

Initial evaluation includes a thorough history and physical exam, diagnostic workup, and a multidisciplinary tumor board discussion with expert radiology and pathology review. Recommended imaging includes computed tomography (CT) chest/abdomen/pelvis for all patients, pelvic magnetic resonance imaging (MRI) for rectal cancers, and positron emission tomography (PET-CT) as indicated. Of note, PET-CT and diffusion-weighted MRI may better characterize peritoneal lesions than standard CT although signet ring cell carcinoma cannot be appreciated on PET-CT [28]. Colonoscopy should also be performed in all patients. Patients should be referred to patient support groups, financial support, fertility counseling, psychosocial support, and social work as indicated [29] [30].

Standard molecular analysis should be conducted, which includes assessing for microsatellite instability (MSI), *RAS* mutations (*KRAS, NRAS, and HRAS*), *BRAF* mutations, ERBB2 (formerly *HER2)* status, and tumor mutational burden (TMB) by next-generation sequencing (NGS) [31] [32]. NGS also has the potential to identify sequence variations, such as *ERBB2* amplifications or *NTRK* gene fusions [33].

Germline testing should be considered as indicated [34]. The prognostic significance of ctDNA is an active area of research, but currently its role in preoperative risk stratification is less established than in postoperative surveillance [35, 36].

#### Block 2: Nonoperative Management

**(% Agreement:** Round 1 = 95%, Round 2 = 99%)

Nonoperative management is recommended for patients with poor performance status (PS) and for patients with extensive solid organ or extraperitoneal metastases. For patients with poor PS, the risk of major surgery may outweigh potential benefits. It is important to discuss with patients and caregivers that CRS ± IPCT is a major abdominal surgery with serious postoperative morbidity rates ranging from 15-33% in patients with CRC-PM [37]. The management of malignant gastrointestinal obstruction, often indicating advanced and unresectable disease, is described in a separate guideline by this consortium group (submitted) [38]. Emerging evidence suggests that CRS ± IPCT may benefit patients with limited extraperitoneal disease, but it requires careful consideration [39]. A recent systematic review of 20 studies revealed a mean overall survival of 26.4 months and a 5-year overall survival rate of 25% in patients receiving combined peritoneal and local treatment for peritoneal and limited liver metastases [40]. Our consensus recommends avoidance of major hepatectomy with CRS. Combined CRS with minor hepatectomy could be considered in select patients with limited liver metastases amenable to a CC0 resection and a peritoneal carcinomatosis index (PCI) < 19 amenable to CC0-1 [39]. While no strict cutoff for ‘limited’ liver disease exists, CRS is discouraged in patients with > four metastatic liver foci. Some experts suggest adding three PCI points for every liver metastasis based on anecdotal experience.

Krukenberg tumors are metastatic tumors of the ovarian lining originating mostly from gastrointestinal adenocarcinomas [41]. Although historically deemed a terminal finding in CRC-PM, they do not necessarily preclude CRS. A retrospective study found that 52% of CRC-PM patients undergoing CRS ± IPCT at a single institution had Krukenberg tumors, with no difference in disease-free survival compared to those without ovarian metastases [42]. Our group agreed that the presence of Krukenberg tumors, or their progression or non-response to systemic therapy, is not an absolute contraindication to CRS ± IPCT.

#### Block 3: Preoperative Systemic Therapy

**(% Agreement:** Round 1 = 94%, Round 2 = 98%)

In the initial management of synchronous CRC-PM, systemic therapy is administered for 3-6 months to potentially downstage tumor burden and target systemic micro-metastases. Additionally, radiation therapy can be considered for rectal cancers following multidisciplinary tumor board discussion. Details regarding systemic therapy regimens are summarized in Table 2.

**Table 2.** Systemic Therapy Regimens for Metastatic Colorectal Malignancy with Peritoneal Involvement.

There remains limited evidence regarding optimal systemic therapy in patients with resectable CRC-PM. Of 72 clinical trials for metastatic CRC from 2003-2016, only seven trials reported inclusion of patients with PM [7]. Our systematic review (Table 4, Supplementary Table 1) synthesized findings from 13 observational studies, which reported receipt of systemic therapy for patients undergoing CRS for CRC-PM, with no published randomized controlled trials (RCTs) to date [43–55]. The timing, duration, and the agents used for neoadjuvant chemotherapy varied across studies. Three studies (<200 patients each) suggested a survival advantage of neoadjuvant therapy followed by CRS compared to upfront CRS [45, 46, 55]. Studies from the Peritoneal Surface Oncology Group International (PSOGI) Global Registry, with a sample size of over 2,000 patients, the United States HIPEC (Hyperthermic Intraperitoneal Chemotherapy) Collaborative, and several single institution studies failed to demonstrate a benefit of neoadjuvant therapy [43, 44, 47–54]. Proponents of neoadjuvant therapy argue for its potential to reduce PCI and increase complete cytoreduction rates, although this was not uniformly demonstrated across studies [55].

Two studies sought to identify high-risk subgroups that may definitively benefit from preoperative or perioperative therapy, including lymph node-positive CRC with PM. Kuijpers et al. observed longer overall survival in 55 patients who received any perioperative chemotherapy compared to 16 patients without chemotherapy in conjunction with CRS-HIPEC (median: 30 months vs. 14 months, p = 0.024) [48]. However, this difference was attenuated after adjusting for major postoperative complications, which was higher in the group that did not receive any perioperative chemotherapy. Among patients who received any systemic chemotherapy, there was no survival difference according to the sequence of administration (neoadjuvant-only, adjuvant-only, or perioperative). Sugarbaker et al. found no overall survival difference between 38 patients with neoadjuvant chemotherapy and 35 without (median 2.3 years vs. 2.9 years, p = 0.94) [53]. Yet, a notable benefit was seen in the subset of 11 patients with a complete or near-complete response to neoadjuvant chemotherapy.

Addressing these gaps, the CAIRO6 study is the first RCT evaluating perioperative systemic therapy and CRS-HIPEC versus CRS-HIPEC alone for resectable CRC-PM. The trial’s phase II segment deemed perioperative systemic therapy safe and feasible, with a 38% major pathologic response rate among patients receiving neoadjuvant therapy [37]. Results from the phase III randomized component of the trial are pending [56].

Given the current lack of standardization in selection of systemic regimens, our consortium group constructed a summary table delineating initial and subsequent systemic therapy regimens for CRC-PM (Table 2). In line with NCCN recommendations, first-line treatment generally includes fluoropyrimidines, such as fluorouracil (5-FU) or capecitabine. Fluorouracil is combined with leucovorin (5-FU/LV) to potentiate its cytotoxic inhibitory effects. Oxaliplatin (FOLFOX or CAPOX), irinotecan (FOLFIRI or CAPIRI), or their combinations (FOLFOXIRI or CAPOXIRI) may augment this backbone regimen.

Anti-vascular endothelial growth factor (VEGF) antibodies, such as bevacizumab, may be added to first-line treatment. Anti–epidermal growth factor receptor (EGFR) antibodies (cetuximab and panitumumab) are recommended to be added for pan-*RAS* wild-type (*KRAS* and *NRAS*) and *BRAF* wildtype metastatic CRC [57]. Transcriptional profiling has shown potential to predict response to anti-EGFR antibodies and might be superior to the historical right and left-sided classification [58, 59]. For robust patients, FOLFOXIRI and bevacizumab may be considered to maximize tumor response [60]. FOLFOXIRI and anti-EGFR therapy combinations has been shown to not improve overall survival based on the TRIPLETE study [61]. Further line therapies, such as trifluridine-tipiracil plus bevacizumab may be employed in refractory metastatic CRC [62]. Immunotherapy is recommended as first-line single agent therapy for microsatellite instability-high (MSI-H) or mismatch repair deficient (MMR-D) tumors [63].

Diagnostic laparoscopy is the gold standard for assessing PCI and estimating the ability to achieve a complete cytoreduction. It may be offered at diagnosis and/or after completion of induction chemotherapy to determine candidacy for CRS ± IPCT and is generally reserved for CRC-PM patients with PCI ≤ 19-25 amenable to complete or near-complete cytoreduction (CC0-1 CRS) [39, 64].

#### Block 4: Upfront Cytoreductive Surgery ± Intraperitoneal Chemotherapy

**(% Agreement:** Round 1 = 77%, Round 2 = 88%)

Consideration for upfront CRS ± IPCT is reserved for highly selected patients with a high-performance status, low to moderate PCI, and low expected surgical morbidity, and complete cytoreduction predicted. There is no universally accepted definition of low or moderate PCI and tends to be surgeon- and institution-dependent. Extensive mesenteric deposits, small bowel deposits, or porta hepatis involvement might preclude complete cytoreduction. Patients with poorly differentiated histology, such as signet ring cell histology, should be treated with systemic therapy prior to CRS consideration.

This block had only 76% agreement in the first round, which improved to 87% in the second round after de-emphasizing the treatment pathway and outlining selection criteria. After CRS, patients should receive adjuvant systemic therapy for three to six months followed by active surveillance. This recommendation is supported by observational data regarding upfront resection of isolated synchronous CRC-PM in 393 patients from the Netherlands Cancer Registry [52]. In a propensity score-matched analysis, adjuvant systemic chemotherapy was associated with improved overall survival compared to active surveillance (median 39.2 months vs. 24.8 months, p = 0.006) [52].

Postulated advantages of upfront surgery include avoiding systemic therapy-adverse events and reduced postoperative morbidity. In a multi-institutional French series, preoperative bevacizumab administration was associated with twice the rate of early complications after CRS ± IPCT for CRC-PM [65]. Additionally, patients may experience disease progression while on systemic therapy, rendering them unresectable [10]. In the phase II component of the CAIRO6 trial, half of the patients declined trial participation due to concerns about systemic therapy’s toxic effects and cancer becoming unresectable during neoadjuvant therapy. Despite this, all patients in the neoadjuvant arm proceeded to surgery, with comparable patient-reported outcomes between the study arms (perioperative systemic therapy with CRS-HIPEC and CRS-HIPEC alone) [37, 66]. Results of the phase III trial of CAIRO 6 are pending and may help address uncertainties surrounding upfront CRS.

#### Block 5: Progression Status After Systemic Therapy

**(% Agreement:** Round 1 = 94%, Round 2 = 99%)

Following completion of systemic therapy, patients should be re-evaluated to determine response to systemic therapy via cross-sectional imaging and serum tumor markers. Diagnostic laparoscopy can be considered to re-evaluate PCI. For patients with no evidence of progression of their metastatic CRC, candidacy for near-complete CRS should be determined.

#### Block 6: No Progression After Systemic Therapy with Complete Cytoreduction Predicted

**(% Agreement:** Round 1 = 92%, Round 2 = 96%)

Patients with no progression after systemic therapy and with a complete cytoreduction predicted should proceed with CRS ± IPCT. Although the PRODIGE 7 trial did not demonstrate a survival benefit with oxaliplatin IPCT added to CRS, trial participants in both arms experienced a median overall survival > 44 months, which is higher than historical reports with systemic therapy alone [10]. Notably, over 95% of patients in this trial received systemic therapy with CRS, with 219 (83%) of 265 patients receiving preoperative chemotherapy [64]. After CRS ± IPCT, additional systemic therapy should be considered, with the goal of completing at least a total of six months of systemic therapy. Patients should then be followed with an active surveillance program.

The cornerstone of curative-intent treatment for CRC-PM remains CRS, with the objective of resecting all visible tumor implants within the peritoneal cavity [67]. Diagnostic laparoscopy is essential for evaluating the PCI and determining the patient’s candidacy for complete or near-complete cytoreduction (CC0-1). Factors contributing that reduce the likelihood of complete CRS, such as extensive mesenteric deposits, small bowel deposits, or porta hepatis involvement, must be considered. Minimally invasive approaches for cytoreduction may be employed in selected patients with low PCI [68, 69].

The role of IPCT in treating CRC-PM remains contentious, as highlighted in the 2018 Chicago Consensus Guidelines. Seminal evidence from the PRODIGE 7 trial, comparing CRS with oxaliplatin HIPEC for 30 minutes and CRS alone, failed to show a significant difference in overall survival (median 41.7 months vs. 41.2 months, p = 0.99), leading to our consensus group recommendation against short duration high dose oxaliplatin HIPEC. The optimal drug dosing and duration are still uncertain, with some experts within our group favoring mitomycin C for ≥ 90 minutes, an approach yet to be tested in RCTs. Evidence from trials on prophylactic HIPEC for high-risk CRC cannot be directly applied to CRC-PM management, as they focus on locally advanced tumors without PM, unlike CRS ± IPCT, which is administered with therapeutic intent. Trials have yielded conflicting evidence. HIPECT4 demonstrated improved locoregional recurrence-free survival with surgical resection and prophylactic mitomycin C HIPEC compared to resection alone, while PROPHYLOCHIP and COLOPEC trials did not demonstrate a benefit with prophylactic oxaliplatin HIPEC [70–72]. Emerging evidence from the ICARuS trial sheds light on Early Postoperative Intraperitoneal Chemotherapy (EPIC) as an additional IPCT modality for CRC-PM and appendiceal cancer [73]. With CRC accrual terminated post-negative results from PRODIGE 7, 75 CRC patients were randomized to HIPEC (n = 40) vs. EPIC (n = 35). Three-year progression-free survival (PFS) did not significantly differ between treatment arms (median 7.7 months vs. 8.8 months, p = 0.14). Given the inconclusive evidence regarding IPCT for CRC-PM, decisions on its use should involve shared decision-making between patients and a multidisciplinary team. Details regarding regional therapy regimens are summarized in Table 3.

**Table 3.** Regional Therapy Regimens for Metastatic Colorectal Cancer.

As highlighted in our systematic review (Table 4, Supplementary Table 1), evidence regarding adjuvant systemic chemotherapy following complete CRS for CRC-PM remains equivocal [43–55]. Its role is better established in patients undergoing upfront CRS without neoadjuvant therapy (Block 3), but its role in a perioperative or “sandwich” regimen is more complex. Advocates of adjuvant systemic chemotherapy emphasize its role in preventing distant systemic relapse post-CRS, as these are more responsive to systemic treatments than isolated PM [44]. Notably, completing chemotherapeutic treatment as planned (typically > 6 cycles), as opposed to partial treatment, is a critical prognostic factor for improved survival [48, 54]. Our review also identified two important adjustments needed in studies investigating adjuvant systemic therapies [44, 49]. The first is major postoperative morbidity, which may occur in over 30% of patients undergoing CRS ± IPCT and often precludes timely initiation of adjuvant systemic therapies. The second involves addressing immortal time bias which may occur due to an imbalance in for early postoperative deaths across study cohorts.

**Table 4.** Key Question 1: In patients with CRC-PM undergoing CRS, what are the optimal sequences and regimens of systemic therapy (neoadjuvant, adjuvant, perioperative)?

#### Blocks 7, 8, & 9: Incomplete Cytoreduction Predicted or Complete Cytoreduction Predicted Despite Progression on Systemic Therapy

**(% Agreement (Blocks 7/8/9):** Round 1 = 93% / 82% / 93%, Round 2 = 99% / 93% / 96%)

For patients with no disease progression after systemic therapy and with an incomplete cytoreduction predicted (Block 7), first-line systemic therapy should be resumed. In case of progression on first-line systemic therapy (Blocks 7/8/9), initiating second line therapies is preferred. CRS ± IPCT may only be considered in selected patients amenable to complete cytoreduction (Block 8). Offering best supportive care and appropriate clinical trials are essential, while following an active surveillance protocol and reassessing candidacy for CRS.

An alternative for patients with unresectable PM is Pressurized Intraperitoneal Aerosolized Chemotherapy (PIPAC), which is primarily employed palliatively in patients who are ineligible for CRS ± IPCT. While current evidence highlights its safety and feasibility, further research into its efficacy is warranted [74, 75]. As per current recommendations, PIPAC should be utilized only in a clinical trial setting.

#### Block 10: Recurrence After CRS

**(% Agreement:** Round 1 = 86%, Round 2 = 96%)

Recurrence following CRS ± IPCT occurs in the peritoneum alone in approximately 60% of patients within five years after surgery [76]. It often prompts the need for additional systemic treatments, with repeat CRS ± IPCT being a potential option for select cases. While much of the literature on repeat CRS ± IPCT focuses on appendiceal neoplasms, evidence supports its safety and feasibility in patients with CRC-PM [77–84]. Positive prognostic indicators for repeat CRS ± IPCT include a low PCI upon recurrence, absence of extraperitoneal metastases, a disease-free interval exceeding 12 months, and no disease progression on systemic therapy.

#### Block 11: Surveillance Strategies

**(% Agreement:** Round 1 = 88%, Round 2 = 99%)

Aligning with the NCCN guidelines for metastatic CRC, recommended surveillance includes obtaining a history, physical examination, tumor markers (CEA), and cross-sectional imaging every three to six months for the first two years then every six months for a total of five years [85, 86]. Colonoscopy should be performed within one year after CRS, unless no preoperative colonoscopy was performed, in which case it should be done within three to six months [87]. There has been conflicting evidence on the role of conventional tumor markers in detecting PM recurrence [88, 89].

While ctDNA can be considered for surveillance in metastatic CRC, its role remains uncertain for patients with PM. KQ2a and KQ2b addressed ctDNA as a surveillance tool postoperatively and while receiving systemic therapy, respectively. KQ2a identified seven studies (Table 5, Supplementary Table 2) yielding equivocal results regarding the utility of ctDNA in postoperative surveillance. Challenges include low detection rates and tissue-plasma discordance in PM compared to extraperitoneal metastases. This may be due to a plasma-peritoneal barrier limiting tumor DNA shedding, contrasting with visceral metastatic sites which are well vascularized [90–92]. However, small retrospective series suggest higher diagnostic accuracy for postoperative relapse with ctDNA compared to standard markers [93, 94]. The studies reviewed in KQ2b are not elaborated upon further as none of them described PM-specific results for response to systemic therapies. Three studies aligned with the above hypothesis by highlighting lower ctDNA mutant allele frequencies in patients with peritoneal-only metastases compared to non-peritoneal (e.g. liver) metastases [92, 95, 96]. The inconclusive evidence regarding the utility of ctDNA in monitoring PM precludes any strong recommendations; thus, utilization should be based on provider discretion [97, 98].

**Table 5.** Key Question 2a: In patients with CRC-PM, does plasma-based liquid biopsy offer better sensitivity, specificity, and lead time therapy compared with standard surveillance modalities in detecting recurrence following CRS?

### Colorectal Cancer with Metachronous Peritoneal Metastases Pathway (Figure 2, Table 1c, Table 1d)

Considering the commonalities between the metachronous and synchronous CRC-PM pathways, the aim of the following text focuses on distinct aspects of the metachronous pathway. Where there are commonalities between the synchronous and metachronous pathways, the text recommendations for the synchronous blocks apply to the metachronous blocks as well. Consensus percentages for round 1 and round 2 for the metachronous pathway are outlined in Tables 3 and 4. We defined metachronous metastases by a disease-free interval (i.e. the duration between diagnosis of the primary tumor and PM) of at least six months. Other definitions have been used in the literature, including a shorter disease-free interval of at least three months or the detection of PM during relapse after resection of the primary tumor [99].

A risk stratification schematic was developed based on clinical and pathological features, dichotomized into low-risk or high-risk (Block 3). High-risk features are a disease-free interval less than one-year, positive lymph nodes, high-grade primary tumor, signet ring histology, high PCI (strict cutoff not defined), and a right-sided primary cancer. The Peritoneal Surface Disease Severity Score (PSDSS) can be considered for additional risk stratification [100, 101]. For patients without high-risk disease features, systemic therapy may be initiated and candidacy for CRS ± IPCT may be ascertained following a diagnostic laparoscopy, as outlined in Blocks 4, 5, and 6. For patients with any high-risk disease features, systemic therapy should be offered for three to six months. Further treatment should be guided based on disease response as assessed by repeat cross-sectional imaging, tumor marker assessment, and diagnostic laparoscopy, as outlined in Blocks 7 and 8. Recommendations for the management of recurrence (Block 9) and surveillance (Block 10) are consistent with the synchronous pathway, the latter not being subjected to consensus voting again in the metachronous pathway. Notably, patients with metachronous CRC-PM may experience earlier recurrence after CRS ± IPCT compared to those with synchronous CRC-PM [102].

## DISCUSSION

Herein we report updated results of a modified Delphi consensus on the clinical management of patients with synchronous and metachronous CRC-PM. Our current consensus group was expanded to include surgical oncologists, medical oncologists, radiologists, pathologists, and patient advocates. Consensus was achieved in all seven question blocks after two rounds of review. Four blocks with < 90% consensus in the synchronous pathway and three in the metachronous pathway underwent revisions after the first modified Delphi round, with subsequent improvements in the levels of agreement. The primary area of disagreement in the synchronous pathway was regarding upfront CRS ± IPCT, which was de-emphasized and highlighted as an option in carefully selected patients alone. Other areas of conflict were the utility of IPCT in addition to CRS, management in the setting of progression while on systemic therapy, and the role of ctDNA testing. These were addressed by recommending consideration for the relevant therapeutic and surveillance approaches based on shared decision-making between patients and a multidisciplinary team.

Major limitations of this expert consensus merit discussion. Firstly, the available evidence for our rapid reviews were of low quality and scarce, which precluded more advanced statistical techniques, like meta-analysis, to synthesize evidence from the included studies. Therefore, the consensus methodology was employed to provide guidance regarding matters of equipoise. Secondly, the expert panel consisted primarily of surgical oncologists. We anticipated this bias during the inception of this study and involved leaders in medical oncology, radiation oncology, palliative care, and other disciplines early on to review feedback from the first Delphi round and outline principles of systemic therapy. Lastly, the Delphi consensus entailed voting on blocks rather than individual itemized recommendations, aligning with the original Chicago Consensus framework. While this approach helped mitigate survey fatigue, it may have compromised the granularity of feedback received.

### National Perspectives

The NCCN Colon Cancer Guidelines recommend systemic therapy for colon cancer with non-obstructing synchronous PM. For obstructing or near obstructing disease, the NCCN recommends surgical management of the obstruction (i.e. resection, ostomy, bypass, or stenting) followed by systemic therapy. This aligns with our pathway regarding malignant gastrointestinal obstruction [38, 86]. However, the NCCN does not make any recommendations on the value of CRS ± IPCT, contrasting substantially with our group.

The 2022 American Society of Clinical Oncology (ASCO) guidelines for the treatment of metastatic CRC recommend CRS along with systemic therapy for CRC-PM. In line with our consensus, the ASCO guidelines emphasize the importance of a multidisciplinary tumor board in the management of peritoneal disease and state that CRS should only be performed at PSM centers. Whereas our consortium does not consider extraperitoneal disease to be an absolute contraindication to CRS, the ASCO guidelines do. The ASCO guidelines also recommend against oxaliplatin-based IPCT and reference PRODIGE 7 as justification for this statement. While our consensus discourages oxaliplatin-based IPCT, specifically of short duration, a conditional recommendation for mitomycin-based IPCT was made [70]. The ASCO guidelines do not propose an alternative IPCT regimen [103].

Similar to our consensus, the 2022 American Society of Colon & Rectal Surgeons (ASCRS) Clinical Practice Guidelines for Colon Cancer makes a strong recommendation for CRS ± IPCT for patients with resectable peritoneal disease. Our consensus and the ASCRS Clinical Practice Guidelines also highlight a potential role for PET-CT in staging metastatic colon cancer [104].

### International Perspective

The recently published 2022 PSOGI Consensus on HIPEC Regimens for Peritoneal Malignancies: Colorectal Cancer was an international consensus of 70 expert panelists who responded to ten clinical questions regarding IPCT regimens for CRC-PM. In line with our consensus, the PSOGI consensus gave a conditional recommendation for HIPEC for patients with CRC-PM and recommended against short duration and high dose oxaliplatin. Both also recommended consideration of repeat CRS and IPCT for peritoneal recurrence at greater than one year after the index CRS [105].

The 2023 European Society for Medical Oncology (ESMO) Clinical Practice Guidelines for metastatic CRC recommend complete CRS and state that IPCT should only be offered in the setting of a clinical trial. They highlight the need for ongoing trials using other HIPEC regimens. Guidelines from PSOGI, ASCO, ESMO, and our group stress the importance of multidisciplinary tumor boards and appropriate referral to PSM centers for CRC-PM [103, 105, 106].

Guidelines from the Japanese Society for Cancer of the Colon and Rectum (JSCCR) refer to the ‘P’ classification system, a scoring system for quantifying peritoneal disease like the PCI; P0 represents no PM, P1 refers to PM adjacent to the primary tumor without distant PM, P2 refers to few distant PM, and P3 involves numerous distant PM. The JSCCR recommends CRS for P1 and P2 disease if the resection is not significantly invasive, similar to our recommendations. The JSCCR does not comment on IPCT, recommends systemic therapy for peritoneal recurrence, and does not identify a role for repeat CRS [107].

A 2019 bi-national survey of Australasian Colorectal Surgeons differed critically from our recommendations in questioning the value of CRS ± IPCT and referral to PSM centers for patients with CRC-PM [108]. It is important to highlight differences in the structuring questions between this survey and our consensus, with the former lumping CRS and HIPEC together whereas ours offered flexibility in considering CRS with or without IPCT. Of note, the survey, a 2018 international PSOGI consensus, and our consensus consider Krukenberg tumors as PM and not an absolute contraindication to CRS [108, 109].

### Patient and Caregiver Perspectives

COLONTOWN is an online community of more than 100 private social media groups for patients with colorectal cancer and their caregivers. Four members of the COLONTOWN community, two patients with CRC-PM and two caregivers of patients with CRC-PM, provided their perspectives on managing this disease. They emphasized the importance of 1) clinical trial enrollment, 2) balancing survival and quality of life goals, 3) nurse navigators, 4) supporting mental health, and 5) obtaining input from PSM experts.

The patients and caregivers reported limited options when it comes to finding a clinical trial that offers a lasting impact, let alone a cure, but they noted that current trials do provide patients with reprieve from chemotherapy. One caregiver said, “I would like to see more support for patients and caregivers researching clinical trials.” The other caregiver recounted that her husband has completed two clinical trials with plans to start a third. The patient has had significantly less side effects from these trials compared to from his chemotherapy. One patient stated, “The more we can be involved in clinical trials, the more hope there is that we will find a cure.” There are limited treatment options for patients with CRC-PM, and these respondents have highlighted the need for more clinical trials in CRC-PM.

## Conclusions

This study reported on a modified Delphi consensus for the management of CRC-PM. Building on the 2018 Chicago Consensus Guidelines, pathways for synchronous and metachronous CRC-PM were updated based on the results of this expert consensus. Three systematic rapid reviews highlighted the limited evidence regarding the utility of ctDNA for surveillance and the optimal systemic therapy for patients with CRC-PM undergoing CRS ± IPCT. These questions and other matters of equipoise, such as the role of IPCT in addition to CRS, warrant further investigation as part of the multimodal treatment of CRC-PM.

## Data Availability

All data produced in the present study are available upon reasonable request to the authors

## Supplementary

**Supplementary Figure 1.** PRISMA Flow Diagram for Key Question 1 regarding systemic therapy for patients with CRC-PM undergoing CRS ± IPCT.

**Supplementary Figure 2.** PRISMA Flow Diagram for Key Question 2a regarding plasma-based liquid biopsy for detecting recurrence after CRS ± IPCT for CRC-PM.

**Supplementary Figure 3.** PRISMA Flow Diagram for Key Question 2b regarding plasma-based liquid biopsy for monitoring response to systemic therapies in patients with CRC-PM.

**Supplementary Table 1.** Quality assessment of studies investigating the optimal sequence of systemic therapy for patients with CRC-PM undergoing CRS ± IPCT using the Newcastle-Ottawa Scale.

**Supplementary Table 2.** Quality assessments of studies investigating the role of plasma-based liquid biopsy in detecting recurrence following CRS ± IPCT using the Quality Assessment of Diagnostic Accuracy Studies Version 2 (QUADAS-2).

**Supplementary Table 3.** Search strategy for Key Question 1.

**Supplementary Table 4.** Search strategy for Key Question 2a.

**Supplementary Table 5.** Search strategy for Key Question 2b.

## Declarations

## Ethics approval and consent to participate

Not applicable.

## Consent for publication

Not applicable.

## Availability of data and materials

The datasets used and/or analyzed during the current study are available from the corresponding author on reasonable request.

## Declaration of Generative AI and AI-assisted technologies in the writing process

During the preparation of this work, the authors used a large language model (ChatGPT V3.5) to revise the manuscript text for coherence and clarity. After using this service, the authors reviewed and edited the content as needed and take full responsibility for the content of the publication.

## Author contributions

KSS, VVB: Ideation and conception, data analysis and interpretation, manuscript writing and editing. MMW, NB, FAG: data interpretation, methodology, manuscript editing. JW, MTW, JTB, EP, SC, LES, DGS: Data analysis and review. CGG: data analysis and review, methodology, project supervision, topic expertise. KPSR, DML, US, APS, JPYCS, CE, MF, JMB: pathway refinement, topic expertise, review and edit of manuscript. KKT: Ideation and conception, experimental implementation, data analysis, review and edit of manuscript, project supervision, funding acquisition, guarantor of project.

## ACKNOWLEDGEMENTS

We thank the Society of Surgical Oncology (SSO) and the Advanced Cancer Therapies program committees for lending our group a dedicated meeting space during their annual conferences. We appreciate the COLONTOWN support group for connecting us with patients and caregivers. We are grateful to the SSO Quality Committee for critically appraising the guideline. We also thank the representatives from the Peritoneal Surface Oncology Group International (PSOGI) for providing perspective commentaries. We appreciate the inputs from Alexandria Brackett, a medical librarian specialist at the Yale Harvey Cushing Library, for examining the rapid review search strategies.

