## Supplementary material for "Consensus Guideline for the Management of Colorectal Cancer with Peritoneal Metastases": Figure 1

### Colorectal Cancer with Synchronous Peritoneal Metastasis

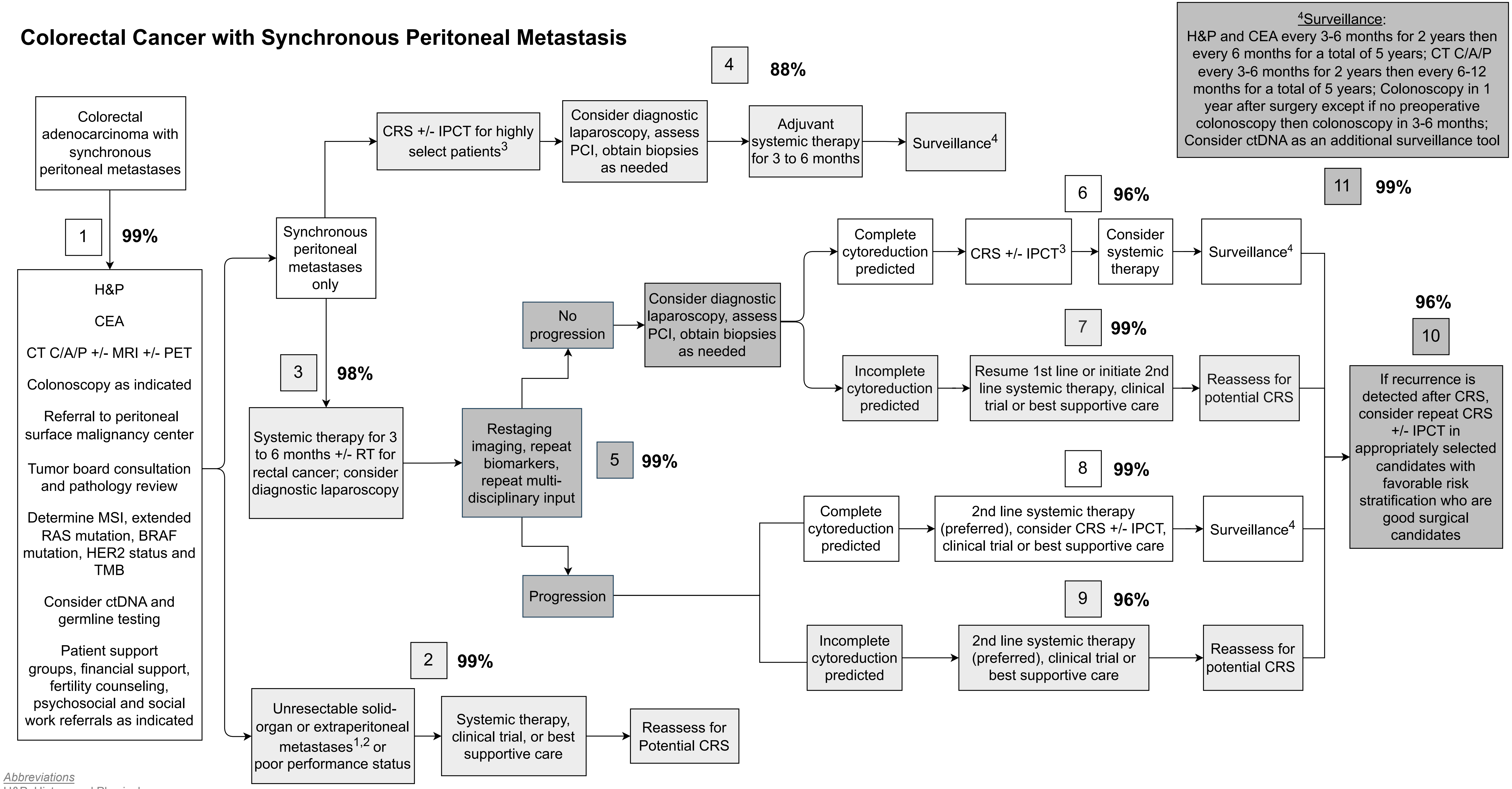


*Abbreviations*
H&P: History and Physical
CEA: Carcinoembryonic antigen
TMB: Tumor mutational burden
IPCT: Intraperitoneal chemotherapy (includes HIPEC and EPIC)
HIPEC: Hyperthermic intraperitoneal chemotherapy
EPIC: Early postoperative intraperitoneal chemotherapy
CRS: Cytoreductive surgery
CT C/A/P: Computed tomography of chest/abdomen/pelvis
MRI: Magnetic resonance imaging
PET-CT: Positron emission tomography-Computed tomography
PCI: Peritoneal cancer index
MSI: Microsatellite instability
ctDNA: circulating tumor DNA
RT: Radiation therapy

[1] Consider palliative procedures for symptomatic patients (ex. bowel obstruction, fistula)

[2] Refer to principles of surgery for patients with resectable liver and/or lung metastases in the context of peritoneal metastasis

[3] Highly select patients: Low PCI, Complete cytoreduction predicted, High performance status, Low expected surgical morbidity

[4] If incomplete CRS, consider systemic therapy, clinical trial, or best supportive care

2023
**Consensus Guidelines**
for Peritoneal Surface Malignancies
