## Supplementary material for "Consensus Guideline for the Management of Colorectal Cancer with Peritoneal Metastases": Figure 2

### Colorectal Cancer with Metachronous Peritoneal Metastasis

[4]Surveillance:
H&P and CEA every 3-6 months for 2 years then every 6 months for a total of 5 years; CT C/A/P every 3-6 months for 2 years then every 6-12 months for a total of 5 years; Colonoscopy in 1 year after surgery except if no preoperative colonoscopy then colonoscopy in 3-6 months; Consider ctDNA as an additional surveillance tool

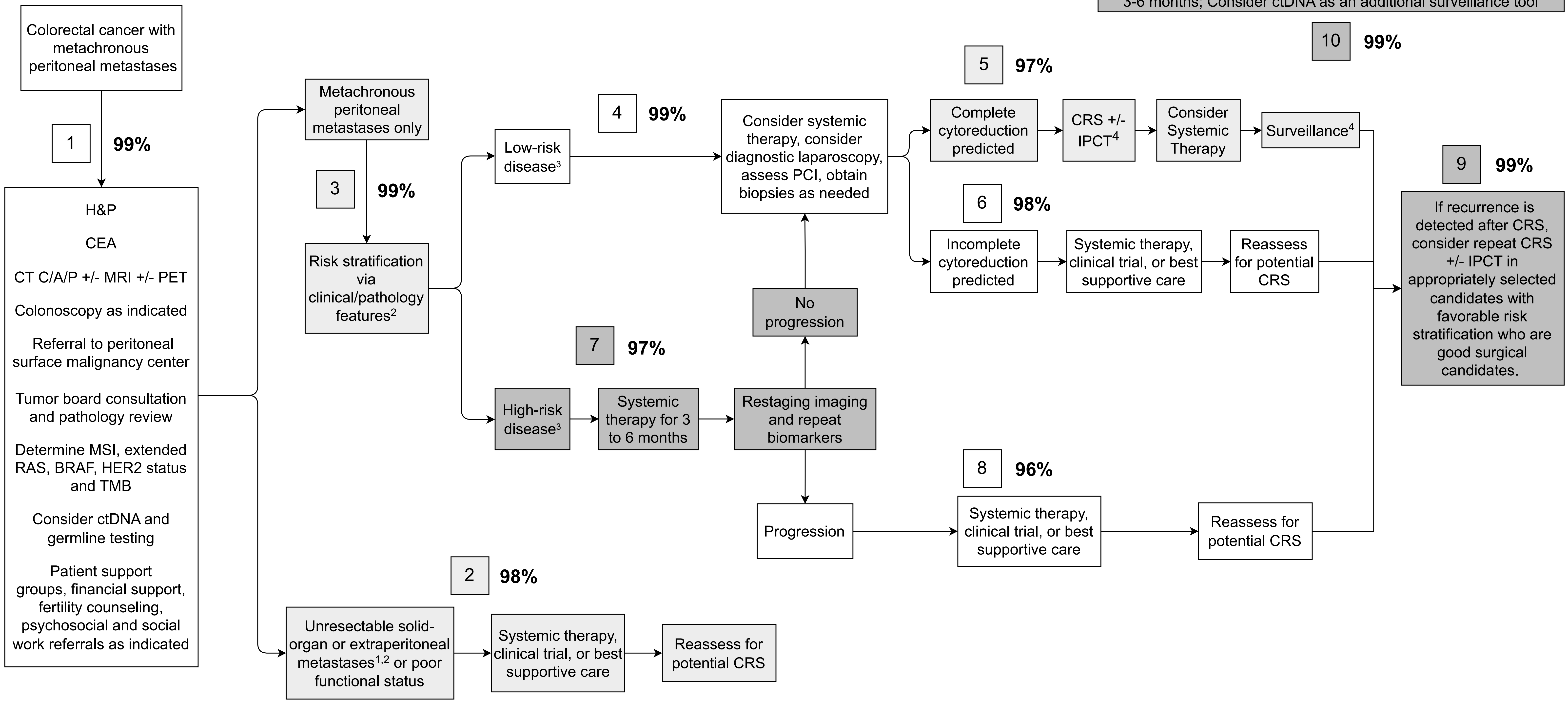


*Abbreviations*
H&P: History and Physical
CEA: Carcinoembryonic antigen
TMB: Tumor mutational burden
IPCT: Intraperitoneal chemotherapy (includes HIPEC and EPIC)
HIPEC: Hyperthermic intraperitoneal chemotherapy
EPIC: Early postoperative intraperitoneal chemotherapy
CRS: Cytoreductive surgery
CT C/A/P: Computed tomography of chest/abdomen/pelvis
MRI: Magnetic resonance imaging
PET-CT: Positron emission tomography-Computed tomography
LN: Lymph node
PSDSS: Peritoneal Surface Disease Severity Score
MSI: Microsatellite instability
ctDNA: circulating tumor DNA

[1]Consider palliative procedures for symptomatic patients (ex. bowel obstruction, fistula)

[2]Refer to principles of surgery for patients with resectable liver and/or lung metastases in the context of peritoneal metastasis

[3]Clinical/pathology features:
- Low risk: Disease-free interval >1 yr, LN-, Low Grade, Low PCI, Left-sided primary cancer
- High risk: Disease-free interval < 1yr, LN+, High Grade, Signet Ring, High PCI, Right-sided primary cancer
- Consider PSDSS for Risk Stratification

[4]If incomplete CRS, consider systemic therapy, clinical trial, or best supportive care

2023
**Consensus Guidelines**
for Peritoneal Surface Malignancies
