## Supplementary material for "Consensus Guideline for the Management of Colorectal Cancer with Peritoneal Metastases": Tables 1-5

**Table 1. Modified Delphi Agreement Table**. Round 1 and Round 2 for Colorectal Cancer with

Synchronous and Metachronous Peritoneal Metastases. Percent agreement includes agree and

strongly agree.

**Table 1a.** Round 1 Agreement Table for Colorectal Cancer with Synchronous Peritoneal Metastases.

|  | **Strongly agree** | **Agree** | **Neither agree nor disagree** | **Disagree** | **Strongly disagree** | **Total** | **% Agree** |
| --- | --- | --- | --- | --- | --- | --- | --- |
| **Block 1** | 106 | 36 | 2 | 0 | 1 | 145 | 98% |
| **Block 2** | 111 | 27 | 5 | 2 | 0 | 145 | 95% |
| **Block 3** | 94 | 43 | 6 | 2 | 0 | 145 | 94% |
| **Block 4** | 67 | 45 | 20 | 13 | 0 | 145 | 77% |
| **Block 5** | 95 | 42 | 6 | 2 | 0 | 145 | 94% |
| **Block 6** | 90 | 44 | 8 | 3 | 0 | 145 | 92% |
| **Block 7** | 86 | 49 | 9 | 1 | 0 | 145 | 93% |
| **Block 8** | 69 | 50 | 17 | 9 | 0 | 145 | 82% |
| **Block 9** | 90 | 45 | 5 | 5 | 0 | 145 | 93% |
| **Block 10** | 76 | 49 | 15 | 5 | 0 | 145 | 86% |
| **Block 11** | 77 | 50 | 12 | 5 | 1 | 145 | 88% |

**Table 1b**. Round 2 Agreement Table for Colorectal Cancer with Synchronous Peritoneal Metastases.

|  | **Strongly agree** | **Agree** | **Neither agree nor disagree** | **Disagree** | **Strongly disagree** | **Total** | **% Agree** |
| --- | --- | --- | --- | --- | --- | --- | --- |
| **Block 1** | 123 | 12 | 0 | 1 | 0 | 136 | 99% |
| **Block 2** | 120 | 14 | 1 | 1 | 0 | 136 | 99% |
| **Block 3** | 117 | 16 | 3 | 0 | 0 | 136 | 98% |
| **Block 4** | 95 | 25 | 7 | 5 | 4 | 136 | 88% |
| **Block 5** | 123 | 11 | 1 | 0 | 1 | 136 | 99% |
| **Block 6** | 116 | 14 | 4 | 1 | 1 | 136 | 96% |
| **Block 7** | 122 | 12 | 1 | 1 | 0 | 136 | 99% |
| **Block 8** | 110 | 16 | 5 | 4 | 1 | 136 | 93% |
| **Block 9** | 117 | 14 | 3 | 1 | 1 | 136 | 96% |
| **Block 10** | 118 | 12 | 4 | 1 | 1 | 136 | 96% |
| **Block 11** | 119 | 15 | 1 | 0 | 1 | 136 | 99% |

**Table 1c**. Round 1 Agreement Table for Colorectal Cancer with Metachronous Peritoneal Metastases.*

|  | **Strongly agree** | **Agree** | **Neither agree nor disagree** | **Disagree** | **Strongly disagree** | **Total** | **% Agree** |
| --- | --- | --- | --- | --- | --- | --- | --- |
| **Block 1** | 102 | 39 | 3 | 0 | 1 | 145 | 97% |
| **Block 2** | 103 | 36 | 4 | 2 | 0 | 145 | 96% |
| **Block 3** | 93 | 43 | 7 | 2 | 0 | 145 | 94% |
| **Block 4** | 84 | 44 | 15 | 1 | 1 | 145 | 88% |
| **Block 5** | 93 | 41 | 6 | 5 | 0 | 145 | 92% |
| **Block 6** | 95 | 42 | 8 | 0 | 0 | 145 | 94% |
| **Block 7** | 98 | 35 | 10 | 2 | 0 | 145 | 92% |
| **Block 8** | 84 | 46 | 11 | 3 | 1 | 145 | 90% |
| **Block 9** | 80 | 45 | 14 | 5 | 1 | 145 | 86% |

**Block 10 was not subjected to consensus voting in the metachronous pathway as it was identical*

*to Block 11 in the synchronous pathway.*

**Table 1d**. Round 2 Agreement Table for Colorectal Cancer with Metachronous Peritoneal

Metastases.*

|  | **Strongly agree** | **Agree** | **Neither agree nor disagree** | **Disagree** | **Strongly disagree** | **Total** | **% Agree** |
| --- | --- | --- | --- | --- | --- | --- | --- |
| **Block 1** | 123 | 12 | 0 | 1 | 0 | 136 | 99% |
| **Block 2** | 120 | 13 | 2 | 1 | 0 | 136 | 98% |
| **Block 3** | 124 | 10 | 1 | 1 | 0 | 136 | 99% |
| **Block 4** | 118 | 17 | 1 | 0 | 0 | 136 | 99% |
| **Block 5** | 117 | 15 | 3 | 1 | 0 | 136 | 97% |
| **Block 6** | 124 | 9 | 3 | 0 | 0 | 136 | 98% |
| **Block 7** | 119 | 13 | 4 | 0 | 0 | 136 | 97% |
| **Block 8** | 117 | 13 | 3 | 2 | 1 | 136 | 96% |
| **Block 9** | 119 | 15 | 1 | 0 | 1 | 136 | 99% |

**Block 10 was not subjected to consensus voting in the metachronous pathway as it was identical*

*to Block 11 in the synchronous pathway.*

**Table 2.** Systemic Therapy Regimens for Metastatic Colorectal Malignancy with Peritoneal Involvement.

| Type of CRC | Stage of therapy | Initial therapy | Subsequent therapy |
| --- | --- | --- | --- |
| Initially unresectable pMMR/MSS mCRC, left-sided, RAS wildtype | Definitive/conversion chemotherapy | FOLFOX or FOLFIRI doublet chemotherapy ± anti-EGFR or anti-VEGF preferred  *FOLFOXIRI triplet chemotherapy (up to 12 cycles) ± anti-VEGF may be considered followed by maintenance 5-FU/leucovorin/bevacizumab | *Regimens as described at left not previously attempted* |
| Other initially unresectable pMMR/MSS mCRC | Definitive/conversion chemotherapy | FOLFOX or FOLFIRI doublet chemotherapy ± anti-VEGF preferred  *FOLFOXIRI triplet chemotherapy (up to 12 cycles) ± anti-VEGF may be considered followed by maintenance 5-FU/leucovorin/bevacizumab | *Regimens as described at left not previously attempted* |
| Complete cytoreduction predicted pMMR/MSS mCRC, left-sided, RAS wildtype | Neoadjuvant chemotherapy | FOLFOX or FOLFIRI doublet chemotherapy ± anti-EGFR or anti-VEGF preferred  *FOLFOXIRI triplet chemotherapy (up to 12 cycles) ± anti-VEGF may be considered followed by maintenance 5-FU/leucovorin/bevacizumab | *Regimens as described at left not previously attempted* |
| Complete cytoreduction predicted pMMR/MSS mCRC | Neoadjuvant chemotherapy | FOLFOX or FOLFIRI doublet chemotherapy ± anti-VEGF preferred  *FOLFOXIRI triplet chemotherapy (up to 12 cycles) ± anti-VEGF may be considered followed by maintenance 5-FU/leucovorin/bevacizumab | *Regimens as described at left not previously attempted* |
| dMMR/MSI-H mCRC | Neoadjuvant/adjuvant chemotherapy | Anti-PD1 ± anti-CTLA-4 or *systemic chemotherapy as recommended above* | Anti-PD1 ± anti-CTLA-4 if no IO given as first line |
| BRAF V600E mCRC | Neoadjuvant/adjuvant chemotherapy | *Systemic chemotherapy as recommended above* | Anti-BRAF + anti-EGFR |
| HER2 | Neoadjuvant/adjuvant chemotherapy | *Systemic chemotherapy as recommended above* | Anti-HER2 therapy |

*Adverse events are more common with triplet chemotherapy. *Abbreviations: mCRC = Metastatic colorectal cancer; IO = immunotherapy; pMMR/dMMR = Proficient mismatch repair/Deficient mismatch repair; MSS/MSI-H = Microsatellite stable/Microsatellite instability – high.*

**Table 3.** Regional Therapy Regimens for Metastatic Colorectal Cancer

with Peritoneal Involvement.

| Regional Regimens | Currently in use/under investigation |
| --- | --- |
| HIPEC | Mitomycin C (preferred)  Oxaliplatin |
| PIPAC | Multiple anti-neoplastic agents are being tested |
| EPIC | Floxuridine |
| IP | Cellular therapies, immunotherapy, STING agonists |

*Abbreviations: HIPEC = Hyperthermic (heated) intraperitoneal chemotherapy;*

*PIPAC = Pressurized intraperitoneal aerosolized chemotherapy; EPIC = Early*

*postoperative intraperitoneal chemotherapy; IP = Intraperitoneal; STING =*

*Stimulation of Transcription factor-1-Interacting Protein 5.*

| First Author, Year  (Country) | Population | Systemic Therapy Regimens | Systemic Therapy Sequence; Duration | Comparison | Sample Size | Peritoneal Cancer Index (PCI, median); Completeness of cytoreduction (CC) or Residual tumor classification (R) | Follow-up Duration | Overall Survival (months); Hazard ratio (95% Confidence Interval) | Major Adverse Events* |
| --- | --- | --- | --- | --- | --- | --- | --- | --- | --- |
| Beal, 2020^1^  (USA) | Patients with CRC-PM undergoing CRS-HIPEC | FOLFOX, FOLFIRI, bev, capecitabine, XELOX, 5-FU + leucovorin | NAC and/or AC; duration not specified | NAC vs. no NAC (Upfront CRS) | 298 | 12.1; CC0: 74.0%, CC1: 15.8% | NR | NAC 32.7m vs. no NAC 22.0m; Adjusted HR 0.8 (95% CI 0.5-1.2) | 35.9% |
| Cashin, 2023^2^  (International) | Patients with CRC-PM undergoing CRS-HIPEC | FOLFOX, FOLFIRI, bev, capecitabine, XELOX, 5-FU + leucovorin | NAC and/or AC; duration not specified | NAC vs. no NAC (upfront CRS),  AC vs. no AC;  Propensity score matching used | 2,093 | 10.1; CC0: 93%, CC1: 5% | 10 years | NAC 34.7m vs. no NAC 37.0m; HR 1.08 (95% CI 0.88-1.32); AC 45.7m vs. no AC 37.0m; HR 0.79 (95% CI 0.64-.97) | 33% |
| Ceelen, 2014^3^  (Belgium) | Patients with CRC-PM undergoing CRS-HIPEC | FOLFOX, FOLFIRI, bev | NAC and/or AC; NAC ≥ 3 months | NAC w/ Bev vs. NAC w/o Bev vs. no NAC | 166 | 4^†^; CC0: 47.6%, CC1: 39.8% | 18 months | NAC w/ Bev 39m vs. NAC w/o Bev 22m vs. no NAC 25m; Adjusted HR NAC w/Bev = 0.31 (95% CI 0.12-0.83) | 35% |
| Devilee, 2016^4^  (Netherlands) | Patients with CRC-PM undergoing CRS-HIPEC | Capecitabine, CAPOX, CAPOX + bev, FOLFOX | NAC or AC; duration not specified | NAC vs. AC | 91 | 6; CC0: 96%, CC1: 4% | 28 months | NAC not reached vs. AC 38.6m; Adjusted HR 0.23 (95% CI 0.07-075) | 18.7% |
| Hanna, 2023^5^  (USA) | Patients with CRC-PM undergoing CRS-HIPEC | FOLFOX ± bev, FOLFIRI ± bev, CAPOX | NAC 6 months or NAC + AC (Sandwich); 6 months | NAC vs. Sandwich | 79 | 11.4; CC0: 85.3%, CC1: 8.8% | NR | NAC 77m vs. Sandwich 61m; Adjusted HR 0.96 (95% CI 0.45-1.32) | NR |
| Kuijpers, 2014^6^  (Netherlands) | Lymph node-positive CRC patients with PM undergoing CRS-HIPEC | FOLFOX, FOLFIRI, bev, capecitabine, XELOX, 5-FU + leucovorin | NAC and/or AC; duration not specified | Any periop chemo vs. no chemo | 73 | 5^†^; ^R1: 87%, R2a: 13% | 47 months | Any chemo 30m vs. no chemo 14m^#^; No significant differences based on chemo sequence (NAC/AC) | 30.1% |
| Maillet, 2016^7^  (France) | Patients with isolated CRC-PM undergoing CRS-HIPEC | FOLFOX, FOLFIRI, bev, cetuximab | NAC and/or AC; duration not specified | AC vs. no AC | 221 | NR; CC0: 100% | 34 months | AC 49 m vs. no AC 43 months; Adjusted HR 1.13 (95% CI 0.7-1.84) | 44.8% |
| Noda, 2023^8^  (Japan) | Patients with CRC-PM undergoing CRS-HIPEC | 5-FU-based ± oxaliplatin ± irinotecan | NAC and/or AC; duration not specified | AC vs. no AC | 123 | NR; R0: 26%, R1: 13.8%, R2: 59.3% | NR | 5-year OS rate R0/1 subgroup = AC 48.2% vs. no AC 38.1%; Adjusted HR 0.366 (95% CI 0.137-0.997) | 21.1% |
| Repullo, 2021^9^  (Belgium) | Patients with CRC-PM with PCI<25 undergoing CRS-HIPEC | FOLFOX or FOLFIRI ± cetuximab or bev | Periop - within 3 months pre/post CRS; ≥ 5 cycles | Periop chemo vs. no chemo | 125 | 6; R0/R1: 100% | 54 months | Chemo 43m vs. no chemo 72m; Adjusted HR 1.46 (95% CI 0.87-2.47) | 21.6% |
| Sugarbaker, 2022^10^  (USA) | Lymph node-positive CRC patients with isolated PM undergoing CRS-HIPEC/EPIC | Not specified | NAC and/or AC; duration not specified | NAC vs. no NAC (Upfront CRS) | 73 | 13; CC0/CC1: 100% | NR | NAC 2.3y vs no NAC 2.9y; HR 1.00 (95% CI 0.62-1.68) | 33.4% |
| Van Eden, 2017^11^  (Netherlands) | Patients with CRC-PM undergoing CRS-HIPEC | CAPOX or FOLFOX or not specified | NAC within 4 months/periop; AC within 3 months | NAC/periop vs. AC vs. chemo only before PC diagnosis (earlier chemo) | 280 | Range: 0-7^†^; R0/R1: 91%, R2a: 8.1% | 29.8 months | NAC/periop 36.9m vs. AC 43.1m vs. earlier chemo 34.0m; Adjusted HR NAC/periop vs. AC 0.84 (95% CI 0.53-1.35) | 30.0% |
| Zhou, 2021^12^  (China) | Patients with CRC-PM undergoing CRS-HIPEC | XELOX or FOLFOX or FOLFIRI ± bev, 5-FU + leucovorin | NAC > 3 cycles &/or AC; duration not specified | NAC vs. no NAC (Upfront surgery) | 52 | 11.9; CC0/CC1: 59.6%, CC2/3: 40.4% | 18.5 months | 2-year OS rate = NAC 67.4% vs. no NAC 32.2%; Adjusted HR 0.55 (95% CI 0.22-1.39) | 34.6% |
| Rovers, 2020^13^  (Netherlands) | Patients with isolated CRC-PM undergoing CRS-HIPEC | CAPOX or FOLFOX or 5-FU or capecitabine or not specified | No NAC (Upfront CRS) ± AC; AC within 3 months | AC vs. no AC (active surveillance); Propensity score matching used | 393 | NR; CC0/CC1: 100% | 25.9 months | AC 39.2m VS. no AC 24.8m; HR 0.66 (95% CI 0.49-0.88) |  |

**Table 4.** Key Question 1: In patients with CRC-PM undergoing CRS, what are the optimal sequences and regimens of systemic therapy (neoadjuvant, adjuvant, perioperative)?

**Major adverse events = Defined variably across studies between Clavien-Dindo/CTCAE 2 through 5; ^†^Uses regional score and not PCI; ^R1 = No residual macroscopic tumor, R2a = Macroscopic residual tumor* $<$*2.5 mm; ^#^Difference attenuated when accounting for major postoperative complications (associated with reduced likelihood of receiving AC). Abbreviations: NR = not reported; CRC-PM = colorectal cancer with peritoneal metastases; CRLM = colorectal liver metastases; CRS = cytoreductive surgery; HIPEC = heated intraperitoneal chemotherapy; CC = completeness of cytoreduction; R = residual tumor classification; Bev = bevacizumab; 5-FU = 5-fluorouracil; ctDNA = circulating tumor DNA; cfDNA = cell-free DNA; PCI = peritoneal cancer index; NAC = neoadjuvant chemotherapy; AC = adjuvant chemotherapy; OS = overall survival; Sandwich = neoadjuvant and adjuvant chemotherapy; HR = Hazard ratio; CI = Confidence interval; Periop = Perioperative; Chemo = Chemotherapy.*

**Table 5.** Key Question 2a: In patients with CRC-PM, does plasma-based liquid biopsy offer better sensitivity, specificity, and lead time therapy compared with standard surveillance modalities in detecting recurrence following CRS?

| **First Author, Year (Country)** | **Population** | **Index Test** | **Index Test Timing** | **Sample Size** | **PM** | **OUTCOMES** | | | |
| --- | --- | --- | --- | --- | --- | --- | --- | --- | --- |
|  |  |  |  |  |  | **Preoperative ctDNA** | **Postoperative ctDNA** | **Sites or Recurrence** | **ctDNA vs. CEA** |
| Beagan, 2020^14^ (Netherlands) | CRC-PM ± limited LMs | Tumor-informed cfDNA | Preop and ≥ 1 postop, then every 3 months for 2 years | 30  (24 CRS-HIPEC) | 100% | Detectable in 33% (8/24) pts; A/w Inferior RFS vs. undetectable ctDNA- HR 3.5 (95% CI 1.1-10.4) | Available for 19 pts - Sn 38% (5/13) and Sp 100% (6/6) for recurrence | Lower Sn of ctDNA for locoregional vs. systemic recurrence (1/8 vs. 4/5) | NR |
| ^#^Baumgartner, 2018^15^  (USA) | PM (multiple primaries) | Tumor agnostic ctDNA (Guardant) | Between 1-2 weeks preop, no postop | 80  (11 CRC) | 100% | Detectable in 39% (31/80) pts;  *High ctDNA A/w inferior PFS- HR 2.4 (95% CI 1.02-5.45) | NR | NR | NR |
| ^#^Baumgartner, 2020^16^  (USA) | PM (multiple primaries) | Tumor agnostic ctDNA (Guardant) | 1-2 weeks preop and 2-5 weeks postop | 71  (16 CRC) | 100% | Detectable in 39% (28/71) overall pts, 62.3% (10/16) CRC pts;  *High ctDNA A/w inferior PFS- HR 3.0 (95% CI 1.6-6.0) | Detectable in 52% (38) overall pts, 63% (10/16) CRC pts;  *High ctDNA A/w inferior PFS:  HR 2.2 (CI 1.1-4.2) | NR | NR |
| ^#^Dhiman, 2023^17^ (USA) | CRC and high-grade AC with PM | Tumor-informed ctDNA (Signatera) | Every 3 months for one year postop | 33  (13 CRC) | 100% | NR | Rising ctDNA A/w inferior DFS vs. undetectable ctDNA- HR 3.7 (95% CI 1.1-12.7);  Rising ctDNA Sn 85.0% (17/20) and Sp 84.6% (11/13) for recurrence | Systemic recurrence A/w higher ctDNA levels vs. peritoneal-only recurrence (199.3 vs. 0.9 MTM/ml) | ctDNA more Sn than CEA (85% vs. 50%) for recurrence |
| Hofste, 2023^18^ (Netherlands) | Metastatic CRC (multiple sites) | Tumor-informed ctDNA | Preop on the day of surgery and 1 week postop | 53 | 11.30% | Detectable in 81% (43/53) pts | Available for 16 pts - Detectable in 25% (4/16) pts | LM A/w higher preop ctDNA detection rate (84% vs. 33%) and ctDNA levels (125.3 vs. 3.3 MTM/ml) compared to PM | Preop ctDNA levels correlated with tumor burden, CEA levels did not |
| Lopez-Rojo, 2020^19^  (Spain) | KRAS-mutated CRC and AC with PM/risk for PM | ddPCR for KRAS mutations in ctDNA | Preop and 48 hours postop | 11  (7 CRC^) | 55% | Detectable in 71% (5/7) CRC pts - Sn 80% (4/5) and Sp 50% (1/2) for recurrence | Available for 5 CRC pts, detectable in 80% (4) pts – Sn 100% (4/4) and Sp 100% (1/1) for recurrence | NR | NR |
| Loupakis, 2021^20^ (Italy) | Metastatic CRC (multiple sites) | Tumor-informed ctDNA (Signatera) | Within 4 weeks postop & at progression or last follow-up | 112 | 14.20% | NR | ctDNA detection (MRD) in 54% (61/112) of pts – Sn 72% (59/82) and Sp 93% (28/30) for recurrence;  MRD A/w inferior DFS - HR 5.8 (95% CI 3.3-10.0) | NR | MRD A/w inferior DFS, CEA not associated –  HR 1.5 (95% CI 0.8-2.7) |

*^#^Baumgartner 2018, Baumgartner 2020, and Dhiman 2023 do not report CRC-specific outcomes; ^HIPEC indication in 7 CRC pts = carcinomatosis (4) and second look for high-risk CRC (3); *High ctDNA levels = MSVAF ≥ 0.25%. Abbreviations: NR= not reported; CRC = Colorectal cancer; PM = Peritoneal metastases; LM = Liver metastases; cfDNA = cell free DNA; ctDNA = circulating tumor DNA; CEA = Carcinoembryonic antigen; CRS = Cytoreductive surgery; HIPEC = Hyperthermic intraperitoneal chemotherapy; Preop = Preoperatively; Postop = Postoperatively; A/w = Associated with; RFS = Recurrence free survival; PFS = Progression free survival; DFS = Disease free survival; HR = Hazard ratio; CI = Confidence interval; Sn = Sensitivity; Sp = Specificity; MTM = Mean tumor molecules; MSVAF = Maximum somatic variant allele fraction; MRD = Minimal or molecular residual disease; ddPCR = digital droplet Polymerase Chain Reaction.*
