## Supplementary Figures 1-3 for "Consensus Guideline for the Management of Colorectal Cancer with Peritoneal Metastases"

**Included**

**Screening**

**Identification**

**Supplementary Figure 1**: PRISMA Flow Diagram for Key Question 1 regarding systemic therapy for patients with CRC-PM undergoing CRS ± IPCT.

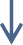

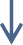

Studies excluded **(n = 102)**

Non-English (n = 5)

Wrong comparator (n = 17) Wrong indication (n = 1) Wrong intervention (n = 18) Wrong study design (n = 43) Sample size insufficient (n = 4)

Wrong patient population (n = 12) Wrong outcome (n = 2)

Studies included in review **(n = 13)**

Studies assessed for eligibility **(n = 115)**

Studies not retrieved **(n = 0)**

Studies sought for retrieval **(n = 115)**

Studies excluded **(n = 1687)**

Studies screened **(n = 1802)**

Studies from databases/registers **(n = 1802)**

MEDLINE (n = 1802)

Included studies ongoing **(n = 0)**

Studies awaiting classification **(n = 0)**

**Supplementary Figure 2**: PRISMA Flow Diagram for Key Question 2a regarding plasma-based liquid biopsy for detecting recurrence after CRS ± IPCT for CRC-PM.

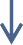

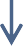

Studies included in review **(n = 7)**

Studies excluded **(n = 92)** Wrong outcomes (n = 1) Wrong intervention (n = 3) Wrong study design (n = 12)

Wrong patient population (n = 76)

Studies assessed for eligibility **(n = 99)**

Studies not retrieved **(n = 0)**

Studies sought for retrieval **(n = 99)**

Studies excluded **(n = 341)**

Studies screened **(n = 440)**

Studies from databases/registers **(n = 440)**

MEDLINE (n = 439)

Citation searching (n = 1)

Included studies ongoing **(n = 0)**

Studies awaiting classification **(n = 0)**

**Included**

**Screening**

**Identification**

**Supplementary Figure 3**: PRISMA Flow Diagram for Key Question 2b regarding plasma-based liquid biopsy for monitoring response to systemic therapies in patients with CRC-PM.

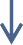

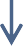

Included studies ongoing **(n = 0)**

Studies awaiting classification **(n = 0)**

**Included**

**Screening**

**Identification**

Studies from databases/registers **(n = 647)**

MEDLINE (n = 647)

References removed **(n = 1)**

Duplicates identified manually (n = 1)

Studies screened **(n = 646)**

Studies excluded **(n = 492)**

Studies sought for retrieval **(n = 154)**

Studies not retrieved **(n = 0)**

Studies assessed for eligibility **(n = 154)**

Studies excluded **(n = 154)** Low sample size (n = 15) Wrong outcomes (n = 24) Wrong comparator (n = 2) Wrong study design (n = 3) No full text available (n = 1)

Wrong patient population (n = 109)

Studies included in review **(n = 0)**
