## Supplementary Tables 1-5 for "Consensus Guideline for the Management of Colorectal Cancer with Peritoneal Metastases"

**Supplementary Table 1.** Quality assessment of studies investigating the optimal sequence of

systemic therapy for patients with CRC-PM undergoing CRS ± IPCT using the Newcastle-Ottawa Scale.^1^

| **Author** | **Selection** | **Comparability** | **Outcomes** | **Overall** |
| --- | --- | --- | --- | --- |
| Beal, 2020 | **** | ** | ** | ******** |
| Cashin, 2023 | **** | ** | *** | ********* |
| Ceelen, 2014 | **** | ** | ** | ******** |
| Devilee, 2016 | *** | ** | ** | ******* |
| Hanna, 2023 | **** | ** | ** | ******** |
| Kuijpers, 2014 | **** | - | *** | ******* |
| Maillet, 2016 | **** | ** | *** | ********* |
| Noda, 2023 | **** | * | ** | ******* |
| Repullo, 2021 | **** | - | *** | ******* |
| Sugarbaker, 2022 | **** | - | *** | ******* |
| Van Eden, 2017 | **** | ** | ** | ******** |
| Zhou, 2021 | **** | ** | ** | ******** |
| Rovers, 2020 | **** | ** | ** | ******** |

*^1^Nine is the maximum number of stars that can be awarded, with six or more stars being*

*considered a good quality study. The maximum number of stars awarded by category is four*

*for Selection, two for Comparability, and three for Outcomes. Abbreviations: CRC-PM = colorectal cancer with peritoneal metastases; CRS = cytoreductive surgery; IPCT = intraperitoneal chemotherapy.*

**Supplementary Table 2**. Quality assessments of studies investigating the role of plasma-based liquid biopsy in detecting recurrence following CRS ± IPCT using the Quality Assessment of Diagnostic Accuracy Studies Version 2 (QUADAS-2).

| **Study** | **RISK OF BIAS** | | | | **APPLICABILITY CONCERNS** | | |
| --- | --- | --- | --- | --- | --- | --- | --- |
|  | PATIENT SELECTION | INDEX TEST | REFERENCE STANDARD | FLOW AND TIMING | PATIENT SELECTION | INDEX TEST | REFERENCE STANDARD |
| Beagan 2020 | ☺ | ☺ | ☺ | ☺ | ☺ | ☺ | ☺ |
| Baumgartner 2018 | ☺ | ? | ? | ? | ☹ | ☹ | ☺ |
| Baumgartner2020 | ☺ | ? | ? | ☺ | ? | ☺ | ☺ |
| Dhiman 2023 | ☹ | ☺ | ☺ | ☺ | ☹ | ☺ | ☺ |
| Hofste 2023 | ☺ | ? | ? | ? | ☹ | ☺ | ☺ |
| Lopez-Rojo 2020 | ☺ | ? | ? | ☹ | ? | ☺ | ☺ |
| Loupakis 2021 | ☺ | ☺ | ☺ | ☺ | ☹ | ☺ | ☺ |

☺ Low Risk	☹ High Risk	?  Unclear Risk

*Abbreviations: CRS = cytoreductive surgery; IPCT = intraperitoneal chemotherapy.*

**Supplementary Table 3.** PubMed search strategy for Key Question 1.

| **Search line** | **Search term** |
| --- | --- |
| **1** | (Neoadjuvant[tw] OR Adjuvant[tw] OR Perioperative[tw]) AND (Chemotherapy OR Immunotherapy[tw] OR Targeted therapy[tw] OR Systemic therapy[tw] OR Drug Therapy[MeSH Major Topic] OR Antineoplastic agents[MeSH Major Topic] OR Drug therapy, Combination[MeSH Major Topic] OR chemotherapy, adjuvant[MeSH Major Topic] OR neoadjuvant therapy[MeSH Major Topic]) |
| **2** | ("surgery peritoneal"[tiab:~5]) OR ("surgery peritoneum"[tiab:~5]) OR ("resection peritoneal"[tiab:~5]) OR ("resection peritoneum"[tiab:~5]) OR (Cytoreduc*[tw] OR CRS[tw]) |
| **3** | Colon cancer[tw] OR Rectal cancer[tw] OR Colorectal cancer[tw] OR Colon neoplasm*[tw] OR Rectal neoplasm*[tw] OR Colorectal neoplasm*[tw] OR Colon tumor*[tw] OR Rectal tumor*[tw] OR Colorectal tumor*[tw] OR (Colorectal neoplasms[MeSH Major Topic]) OR (Colorectal Surgery[MeSH Major Topic]) OR ((colon*[tw] OR rectal*[tw] OR colorectal*[tw]) AND (periton*[tw]) AND (metast* OR carcinomatosis*)) |
| **4** | #1 AND #2 AND #3 |

**Supplementary Table 4.** PubMed search strategy for Key Question 2a.

| **Search line** | **Search term** |
| --- | --- |
| **1** | (Colon cancer OR Rectal cancer OR Colorectal cancer OR Colon neoplasm* OR Rectal neoplasm* OR Colorectal neoplasm* OR Colon tumor* OR Rectal tumor* OR Colorectal tumor* OR (Colorectal neoplasms[MeSH Major Topic]) OR (Colorectal Surgery[MeSH Major Topic]) OR (Gastrointestinal neoplasms[MeSH Major Topic])) |
| **2** | (Surgery[tw] OR Surgeries[tw] OR hepatectom*[tw] OR resect*[tw] OR Cytoreduct*[tw] OR CRS[tw] OR intraperitoneal chemotherapy[tw] OR HIPEC[tw] OR (Hyperthermic Intraperitoneal Chemotherapy[MeSH Major Topic])) |
| **3** | (Cell-Free Nucleic Acids[MeSH Major Topic]) OR (Circulating Tumor DNA[MeSH Major Topic]) OR Liquid biopsy[tw] OR Circulating tumor DNA[tw] OR CtDNA[tw] OR Cell free DNA[tw] OR CfDNA[tw] |
| **4** | #1 AND #2 AND #3 |

**Supplementary Table 5.** PubMed search strategy for Key Question 2b.

| **Search line** | **Search term** |
| --- | --- |
| **1** | (Colon cancer OR Rectal cancer OR Colorectal cancer OR Colon neoplasm* OR Rectal neoplasm* OR Colorectal neoplasm* OR Colon tumor* OR Rectal tumor* OR Colorectal tumor* OR Colon tumour* OR Rectal tumour* OR Colorectal tumour* OR (Colorectal neoplasms[MeSH Major Topic]) OR (Colorectal Surgery[MeSH Major Topic]) OR (Gastrointestinal neoplasms[MeSH Major Topic])) |
| **2** | (Metasta*[tw] OR Periton*[tw] OR (Neoplasm Metastasis[MeSH Major Topic]) OR (Neoplasm, Residual[MeSH Major Topic])) |
| **3** | Neoadjuvant[tw] OR Adjuvant[tw] OR Perioperative[tw] OR Chemotherapy OR Immunotherapy[tw] OR Targeted therapy[tw] OR Systemic therapy[tw] OR Drug*[tw] OR EGFR*[tw] OR Cycle*[tw] OR Challenge*[tw] OR Rechallenge*[tw] OR Respons*[tw] |
| **4** | (Cell-Free Nucleic Acids[MeSH Major Topic]) OR (Circulating Tumor DNA[MeSH Major Topic]) OR Liquid biopsy[tw] OR Circulating tumor DNA[tw] OR CtDNA[tw] OR Cell free DNA[tw] OR CfDNA[tw] |
| **5** | #1 AND #2 AND #3 AND #4 |
